# LncRNA PAN3-AS1 modulates cilium assemble signaling pathway through regulation of RPGR as a potential MS diagnostic biomarker: integrated systems biology investigation

**DOI:** 10.1101/2024.06.03.24307702

**Authors:** Yasaman Mostaghimi, Mohammad Haddadi, Zohreh Hojjati

## Abstract

**Introduction:** Multiple sclerosis (MS), an autoimmune condition of the central nervous system (CNS), can lead to demyelination and axonal degeneration in the brain and spinal cord, which can cause progressive neurologic disability. MS symptoms include autonomic dysfunction, such as insomnia, neuropathic and nociceptive pain, cognitive impairment, spasticity, dysesthesia, anxiety, and depression, significantly reducing patient quality of life. These symptoms appear in addition to the progressive decline in the nervous system’s motor abilities. In this investigation, we performed an integrated bioinformatics and experimental approach to find the expression level and interaction of a novel long non-coding RNA (lncRNA), PAN3-AS1, in MS samples.

**Methods:** Microarray analysis was performed by R Studio using GEOquery and limma packages. lncRNA-mRNA RNA interaction analysis was performed using the lncRRIsearch database. Pathway enrichment analysis was performed by KEGG and Reactome online software through the Enrichr database. Protein-protein interaction analysis was performed by STRING online software. Gene ontology (GO) analysis was performed by Enrichr database.

**Results:** Based on microarray analysis, lncRNA PAN3-AS1 has a significantly low expression in MS samples compared to the control (logFC: −1.2, adj. P. Val: 0.03). qRT-PCR results approved bioinformatics analyses. ROC analysis revealed that PAN3-AS1 could be considered a potential diagnostic biomarker of MS. Based on lncRNA-mRNA interaction analysis, lncRNA PAN3-AS1 regulates the expression level of RPGR. RPGR and its protein interactome regulate the cilium assembly, chaperon-mediated autophagy, and microarray biogenesis.

**Conclusion:** lncRNA PAN3-AS1, as a significant low-expressed lncRNA in MS samples, could be a potential diagnostic MS biomarker. PAN3-AS1 might regulate the expression level of cilium assembly and chaperon-mediated autophagy. Dysregulation of PAN3-AS1 might affect the expression of RPGR and its protein interactome.

## 1. Introduction

Multiple sclerosis (MS) is an autoimmune-related chronic central nervous system inflammatory disease primarily by activated T cells, with emerging evidence of a major impact from B cells and innate immune system cells (Yamout & Alroughani, 2018). Historically, multiple sclerosis has been characterized as an organ-specific T-cell mediated autoimmune illness. However, the success of B-cell targeted treatments calls into question the conventional T-cell autoimmune dogma (Greenfield & Hauser, 2018). Multiple sclerosis is becoming a more worldwide illness (Browne et al., 2014). The latitudinal gradient in MS prevalence is significantly associated with UVB radiation, which enhances the generation of cutaneous vitamin D (vD). Low vD levels, decreased vD consumption, decreased outdoor exercise, and higher MS susceptibility associated with genetic variants generating low vD levels have all been linked to vD being involved in the MS causative pathway (Sintzel et al., 2018).

Previous studies revealed some important regulatory signaling pathways that are related to MS. For example, Vakrakou et al. in 2022 revealed the roles of the mTOR signaling pathway in MS. Many autophagy-inducing circumstances, including food, cellular energy, or growth factor restriction, have been found to suppress mTORC1 activity, implying an inverse link between autophagy induction and mTORC1 activation. mTOR regulates many essential stages in autophagic pathways, including the activation of the beclin-1 complex and LC3-II (Vakrakou et al., 2022). Sphingosine-1-phosphate (S1P) modulation is an intriguing target for the therapy of a variety of autoimmune illnesses. Because of a better knowledge of fingolimod’s mechanism of action, more selective second-generation S1PR modulators have been developed. S1PR subtype 1 (S1PR1) is expressed on the cell surface of lymphocytes, which are known to play an essential role in the pathophysiology of MS. Understanding S1PR1’s involvement aided in the development of pharmaceutical methods focused on this target and theoretically lowered the safety issues associated with fingolimod usage (Pérez-Jeldres et al., 2021). S1PRs were originally targeted as a therapeutic method for multiple sclerosis (MS), and four S1P modulators (fingolimod, siponimod, ozanimod, and ponesimod) are presently licensed for therapy. New S1PR modulators are being tested in clinical trials for MS and other immune-mediated disorders such as inflammatory bowel disease (IBD), rheumatoid arthritis (RA), systemic lupus erythematosus (SLE), and psoriasis (Bravo et al., 2022).

Long non-coding RNAs (lncRNAs) play an important part in complicated biological processes, and their persistence in bodily fluids, along with tissue specificity, makes these macromolecules interesting diagnostic candidates for MS diagnosis (Ghoveud et al., 2020a). Different studies revealed the potential roles of lncRNAs in different diseases. For example, lncRNA BAIAP2-AS1 might have a significant role in the regulation of ADAMTS5 in breast cancer (BC) samples (Tavousi et al., 2022). Based on previous studies, LINC00520 (Ezzati et al., 2022), lncRNA NORAD (Rezvani et al., 2023), and LINC01521 (Zahra et al., 2023) also are the potential BC biomarkers. BAIAP2-AS1 also could regulate ECM receptor signaling pathway in gastric cancer samples (Barani et al., 2022). Based on the study of Shaker et al. in 2019, the expression level of lncRNA MALAT1 has a significantly high expression in the SPMS subgroup of MS patients. However, the expression of MALAT1 has no significant change in the RRMS subgroup (Shaker et al., 2019). Based on the study of Ghoveud et al. in 2020, lncRNA RP11-530C5.1 has a significant up-regulation, and lncRNA AL928742.12 has a significantly low-expression in MS patients. Also, based on this study, the expression of RP11-530C5.1 and AL928742.12 has a significant positive correlation with AWR and IGHA2, respectively, and these results suggest novel MS biomarkers (Ghoveud et al., 2020b). lncRNA PAN3-AS1 is a long non-coding RNA located in chromosome 13. This gene is the antisense sequence of the PAN3 gene. Based on genecards, PAN3 is thought to play a role in the activity of poly(A)-specific ribonuclease. It is anticipated to participate in the shortening of the poly(A) tail of nuclear-transcribed mRNA. The function of PAN3 is likely upstream of or associated with the deadenylation-dependent decapping of nuclear-transcribed mRNA. It is also believed to contribute to the positive regulation of the assembly of cytoplasmic mRNA processing bodies and to be involved in protein targeting. So, PAN3-AS1 could have a regulatory role in the mentioned processes through the regulation of the PAN3 expression level. However, there was no previous study about the regulatory role of PAN3-AS1 in MS.

In this study, we aimed to find a novel dysregulated lncRNA in MS samples and demonstrate a regulatory interaction network based on selected lncRNA. In this expression evaluation, we analyze the expression pattern of lncRNA to find the biomarker capability of selected lncRNA. Another goal of this study is to find the effect of demonstrated regulatory networks on MS-related signaling pathway.

## 2. Materials and Methods

### 2.1. Microarray data analysis

GSE43591 (Jernås et al., 2013)microarray dataset (GPL520 [HG-U133_Plus_2] Affymetrix Human Genome U133 Plus 2.0 Array) analysis was done to identify novel possible MS biomarkers based on expression patterns. In this investigation, limma (Ritchie et al., 2015) was used to analyze 10 MS samples and 10 control samples obtained from Bioconductor. The aforementioned gene expression dataset was obtained from Bioconductor using the GEOquery (Davis & Meltzer, 2007) package and processed with R Studio (4.1.2). The significant thresholds were |logFC| > 0.5 and adj. P. Val < 0.05. Plots of the aforementioned studies were created with the CRAN-available ggplot2 and pheatmap package.

### 2.2. Bioinformatics analysis

lncRNA–mRNA interaction analysis was performed using lncRRIsearch online database (http://rtools.cbrc.jp/LncRRIsearch/) (Fukunaga et al., 2019). Protein – protein interaction analysis was performed by STRING online software (https://string-db.org/) (Jensen et al., 2009). mRNA – miRNA interaction analysis was implemented by miRWalk database (mirwalk.umm.uni-heidelberg.de/) (Dweep & Gretz, 2015; H et al., 2011; Sticht et al., 2018). Pathway enrichment analysis was executed by enrichr (https://maayanlab.cloud/Enrichr/) (Chen et al., 2013; Kuleshov et al., 2016; Xie et al., 2021) and reactome (https://reactome.org/) (A et al., 2018; B et al., 2020; Griss et al., 2020) databases. Visualization of RNA interaction network was performed by cytoscape (3.9.1) (Otasek et al., 2019; Shannon et al., 2003).

### 2.3. Study population

This study was approved by the Ethics Committee of University of Isfahan (Ethics Code: IR.UI.REC.1402.016). All the individuals provided written informed consent letter before contribution to this study. In this experiment, 48 MS blood samples in the two SPMS and RRMS state and 48 blood samples of healthy control individuals were analyzed. The MS patients were in the age range of 31-61 years. The healthy contrals were older than 50 and had no previous MS history.

### 2.4. qRT-PCR experiment

According to the manufacturer’s instructions, total RNA was extracted from cells using RNX_PLUS reagent (CinnaGen company, Tehran, Iran). A RT-PCR kit (ROJE Technologies, Tehran, Iran) was utilized to reverse transcribe total RNA. FOR REAL-TIME QPCR, CFX real-time equipment (Bio-rad, Hercules, CA, USA) and a standard SYBR Green PCR kit (Yekta Tajhiz Azma, Tehran, Iran) were used. The relative expression was computed using the ddCt approach. The GAPDH transcription was used as an internal control.The primers of lncRNA in this study were designed by oligo 7 softwere (version 7) listed in Table1.

### 2.5. Statistical analyses

GraphPad Prism (version 8) was used to do statistical analyses and visualizations of the real-time PCR data. To measure the expression levels, the Ct method was employed to compare the qRT-PCR results from the MS and control samples. The Shapiro-Wilk test was employed to determine if distribution of the expression data was normal. The expression levels in the MS and control samples were compared using the paired t-test and Wilcoxon test on the Ct data. R Studio (4.1.2) was used to conduct the DEG analysis on the microarray data. On the real-time PCR datasets, the GraphPad prism was used to perform receiver operating characteristic (ROC) analysis based on sensitivity and specificity. The significant criterion for this investigation was a p-value less than 0.05. In the ROC analysis, AUC values between 0.7 and 0.8 are regarded acceptable, 0.8 and 0.9 are considered good (indicating a respectable biomarker), and 0.9 and 1 are considered strong (identifying a perfect biomarker).

## 3. Results

### 3.1. lncRNA PAN3-AS1 has a significant low-expression in MS patients

Microarray analysis was performed to find a novel dysregulated lncRNA in the MS samples. Boxplot of the expression data (Figure 1) revealed that all microarray samples had a suitable quality, and there were no noisy samples in this dataset. Based on microarray analysis, among 54613 genes of GSE43591, there were 127 up-regulated and 103 low-expressed genes in MS samples compared to control. Figure 2 shows the top dysregulated genes of this study. lncRNA PAN3-AS1 has a significantly low expression (logFC: −1.2, adj.P.Val: 0.03) in the MS patients as a novel potential diagnostic MS biomarker (Figure 3).

**Figure 1:**
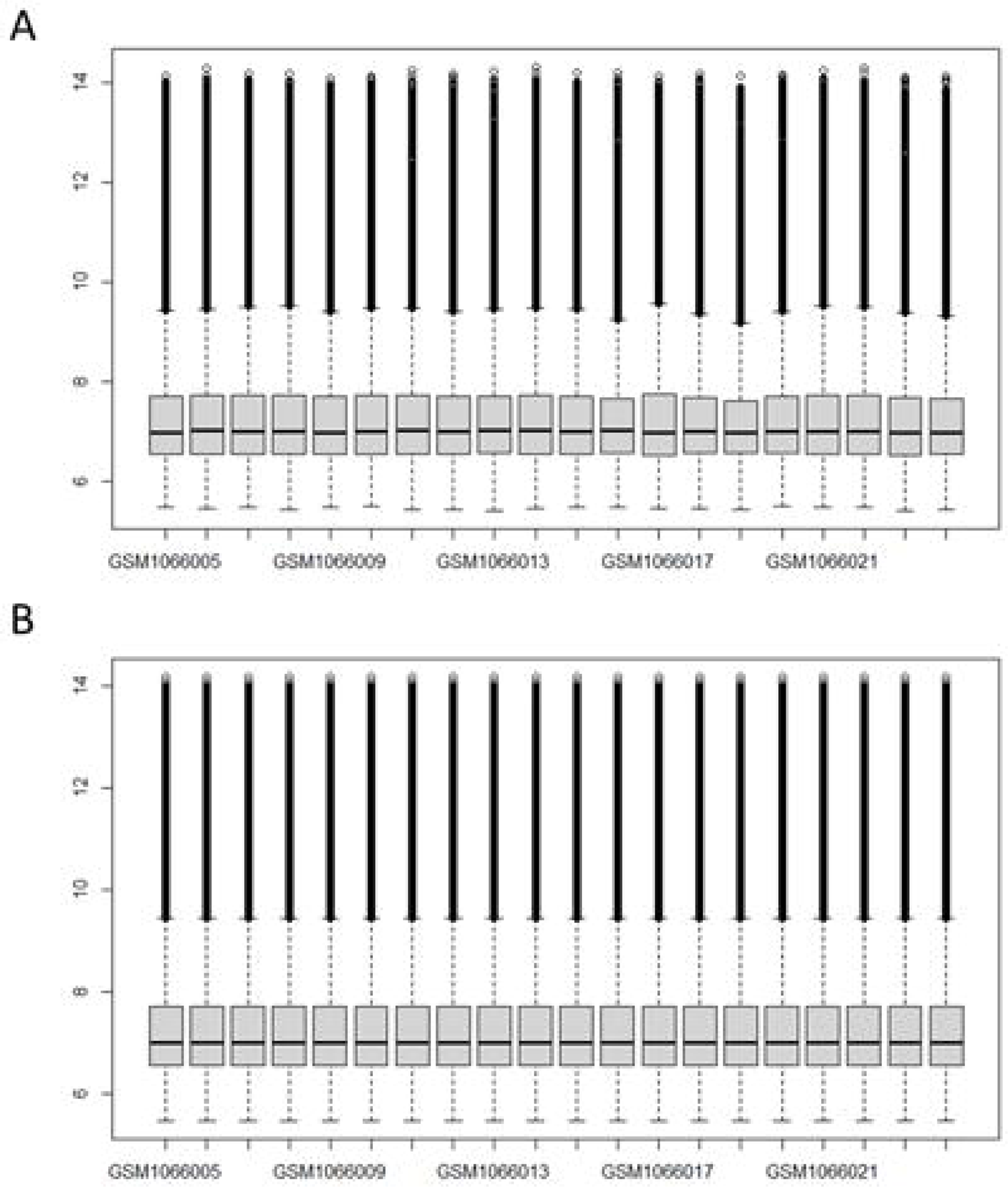
Boxplot of raw (A) and normalized (B) microarray expression dataset (GSE43591). Based on this plot, the median and quartiles of each box (sample) are in a straight line, and this type of boxplot indicates the high quality of microarray samples.

**Figure 2:**
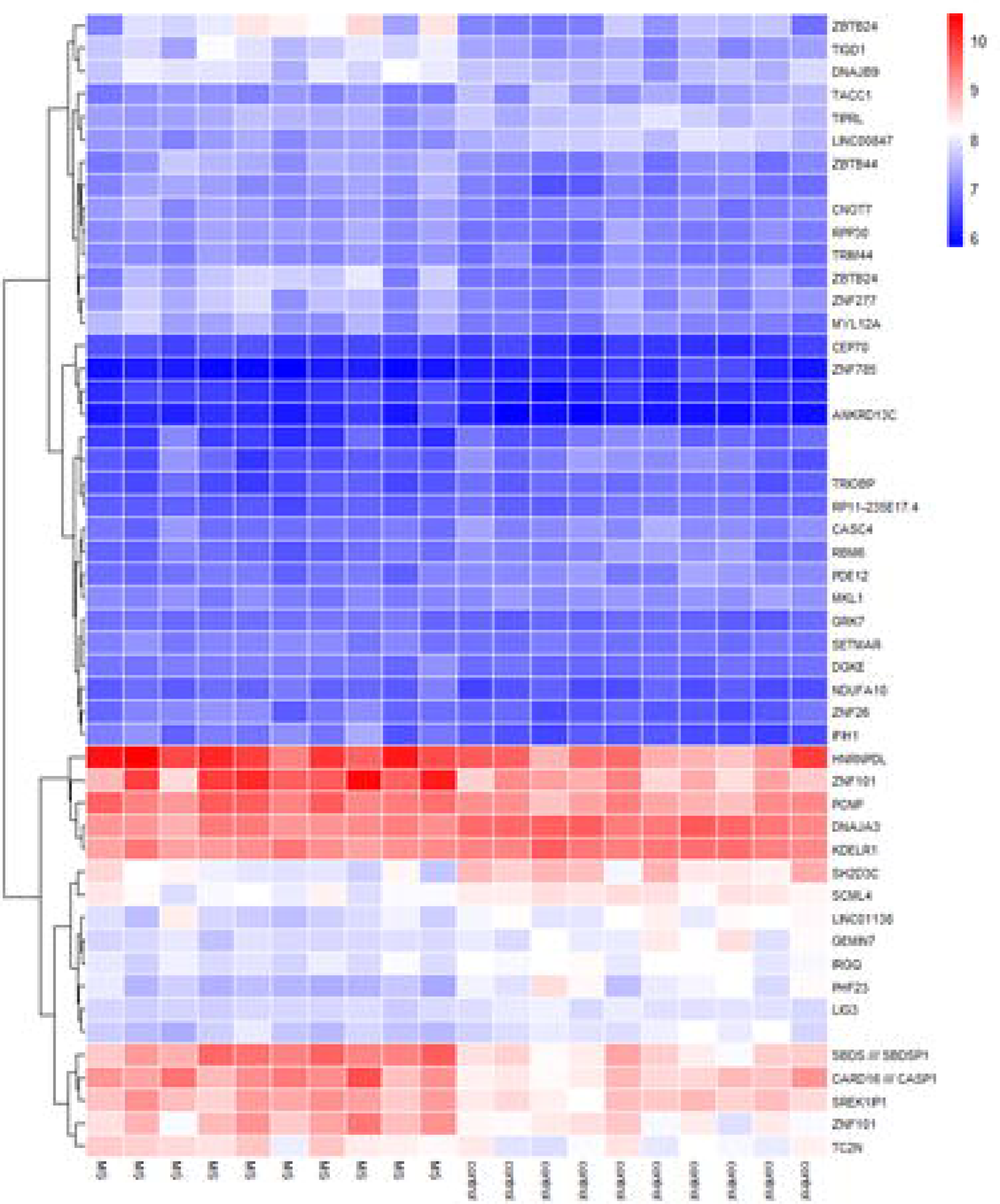
Heatmap of top differentially expressed genes in GSE43591.

**Figure 3:**
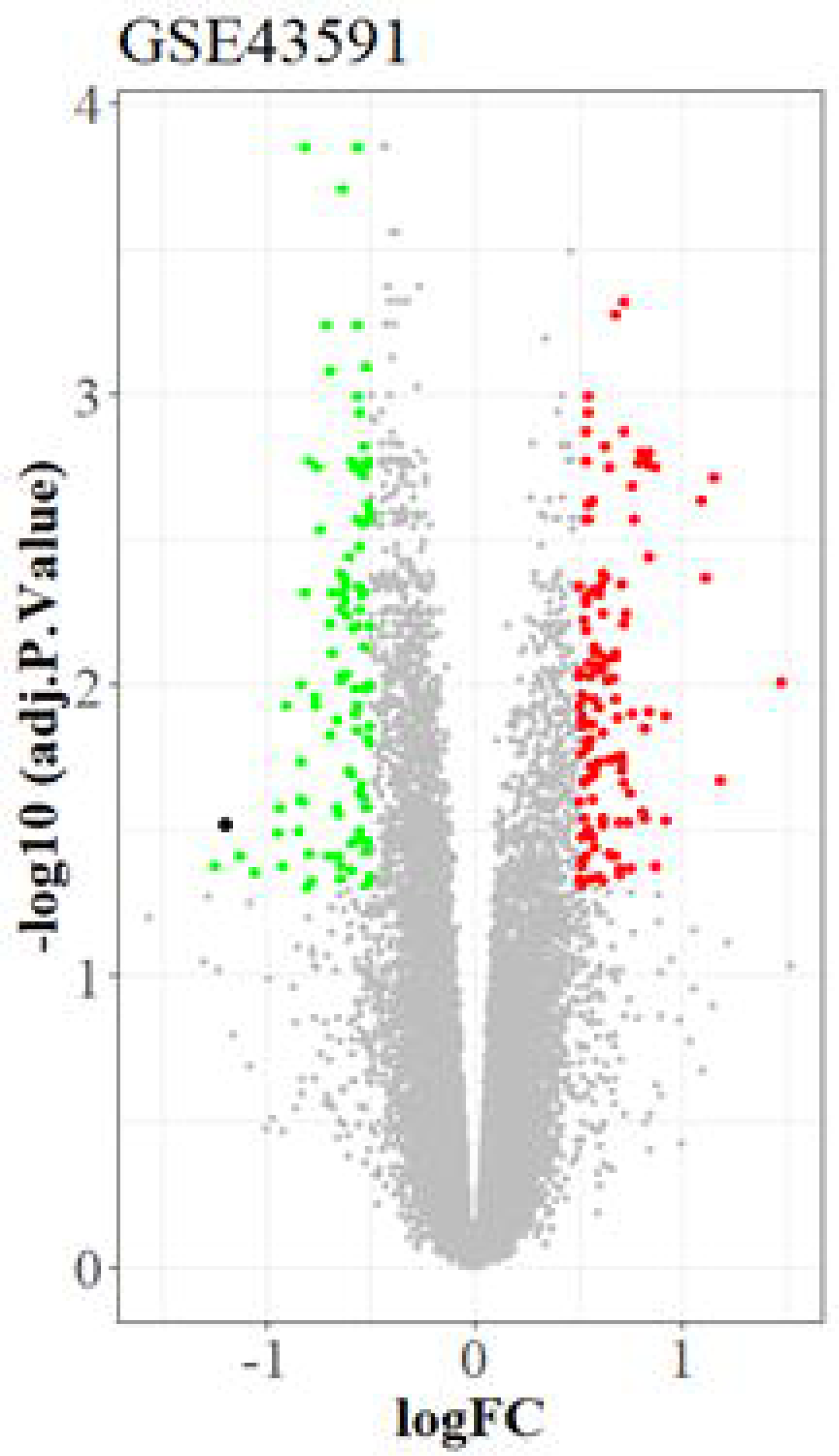
Volcano plot of genes in GSE43591. Up-regulated (red color) and down-regulated (green color) genes are indicated in this plot. PAN3-AS1 is indicated by a black point as a significant low-expressed lncRNA.

A qRT-PCR experiment was performed on 48 MS and 48 normal samples to validate PAN3-AS1 expression based on obtained microarray data analysis reports. Based on the qRT-PCR experiment, PAN3-AS1 has a significant down-regulation in MS samples (logFC; −3.248, p-value < 0.0001). Also, based on ROC analysis, PAN3-AS1 could be considered a potential excellent diagnostic biomarker of MS (AUC: 0.9288, p-value < 0.0001, Figure 4). However, there was no significant correlation between the expression level of PAN3-AS1 with age and different MS subgroups (Figure 5).

**Figure 4:**
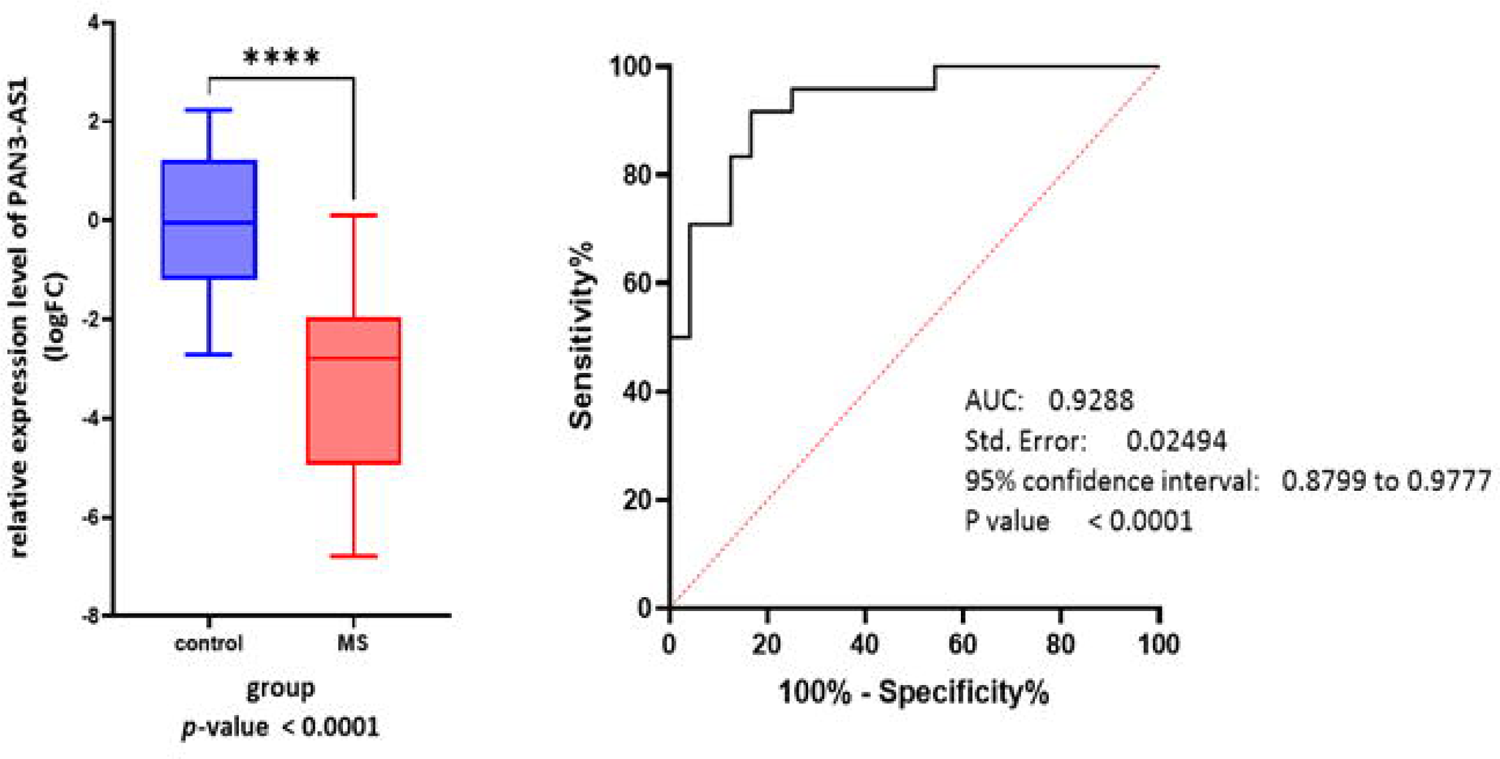
qRT-PCR experiment. Based on the analysis of qRT-PCR data, PAN3-AS1 has a significantly high expression in MS samples. Also, based on ROC analysis, PAN3-AS1 could be considered a potential diagnostic biomarker of MS.

**Figure 5:**
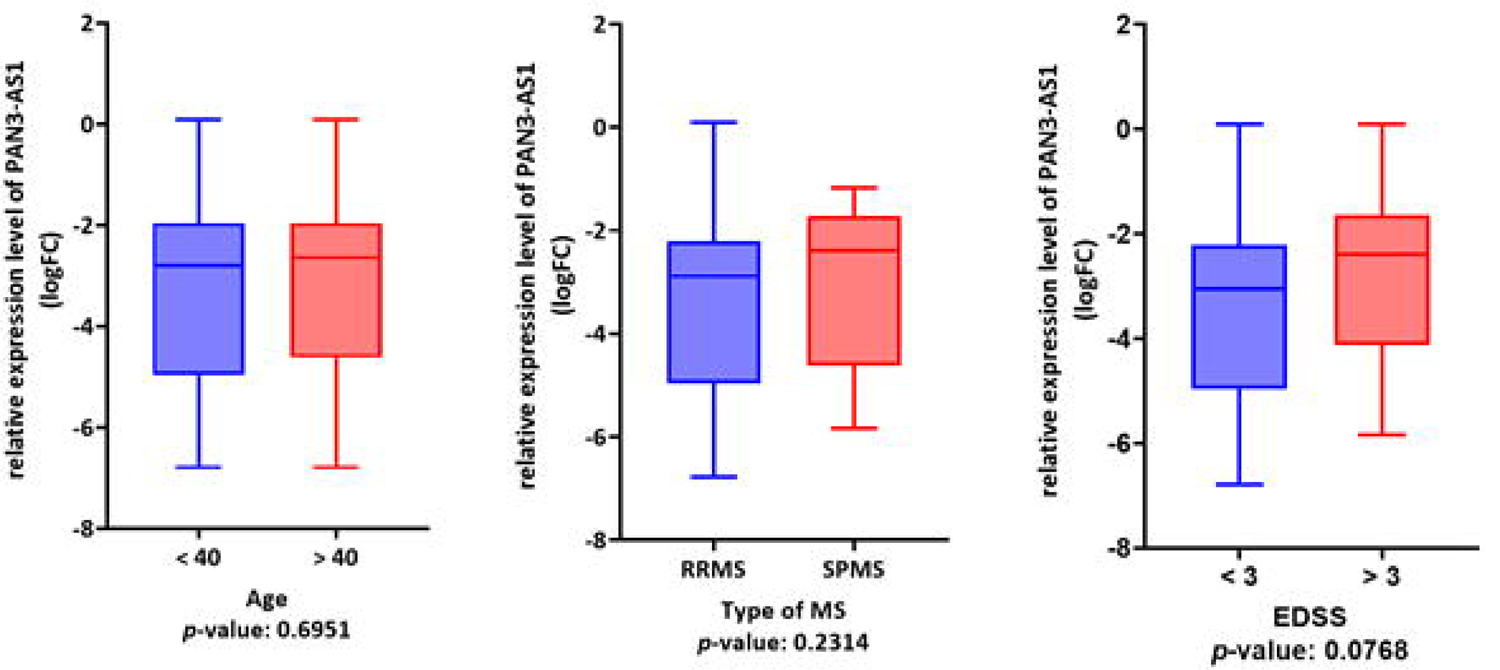
Clinicopathological analysis of PAN3-AS1 expression level based on qRT-PCR data. The expression level of PAN3-AS1 has no significant correlation with age and subgroups of MS patients.

### 3.2. PAN3-AS1 regulates cilium assembly signaling pathway through interaction with RPGR

lncRNA-mRNA interaction analysis revealed the top 10 mRNAs regulated by PAN3-AS1 (Table 2). Based on the mentioned analysis, PAN3-AS1 could significantly regulate mRNA RPGR (sum of energy: −2576.23 kcal/mol). miRNA interaction analysis revealed that hsa-miR-6872-3p has a significant interaction with hsa-miR-939-5p (energy: −29.3, position: 3’UTR, Table 3, Figure 6). Protein-protein interaction analysis revealed three different clusters of proteins that are in the interaction network of RPGR (Table 4, Figure 7).

**Figure 6:**
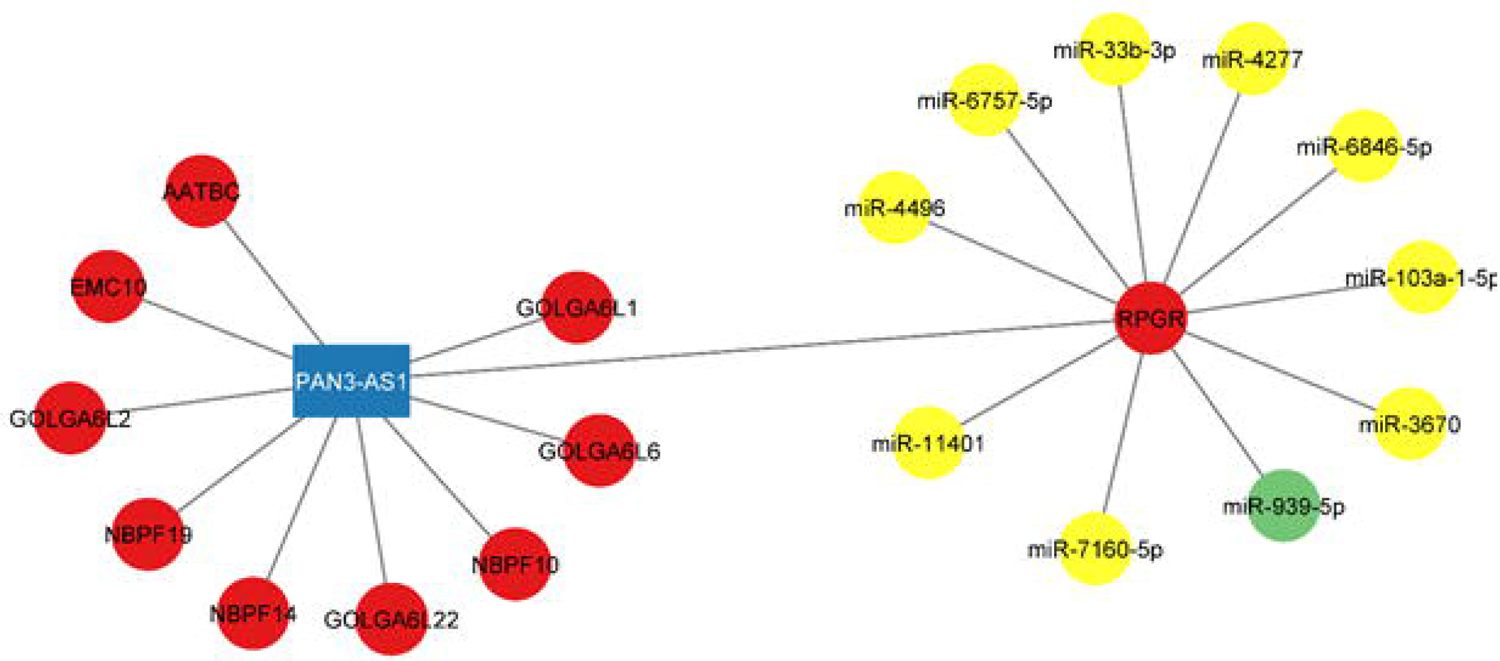
RNA interaction of lncRNA PAN3-AS1 and RPGR. Red nodes indicate mRNAs and yellow nodes indicate miRNAs. miR-939-5p is shown in the plot by the green node as this study’s most important regulatory miRNA.

**Figure 7:**
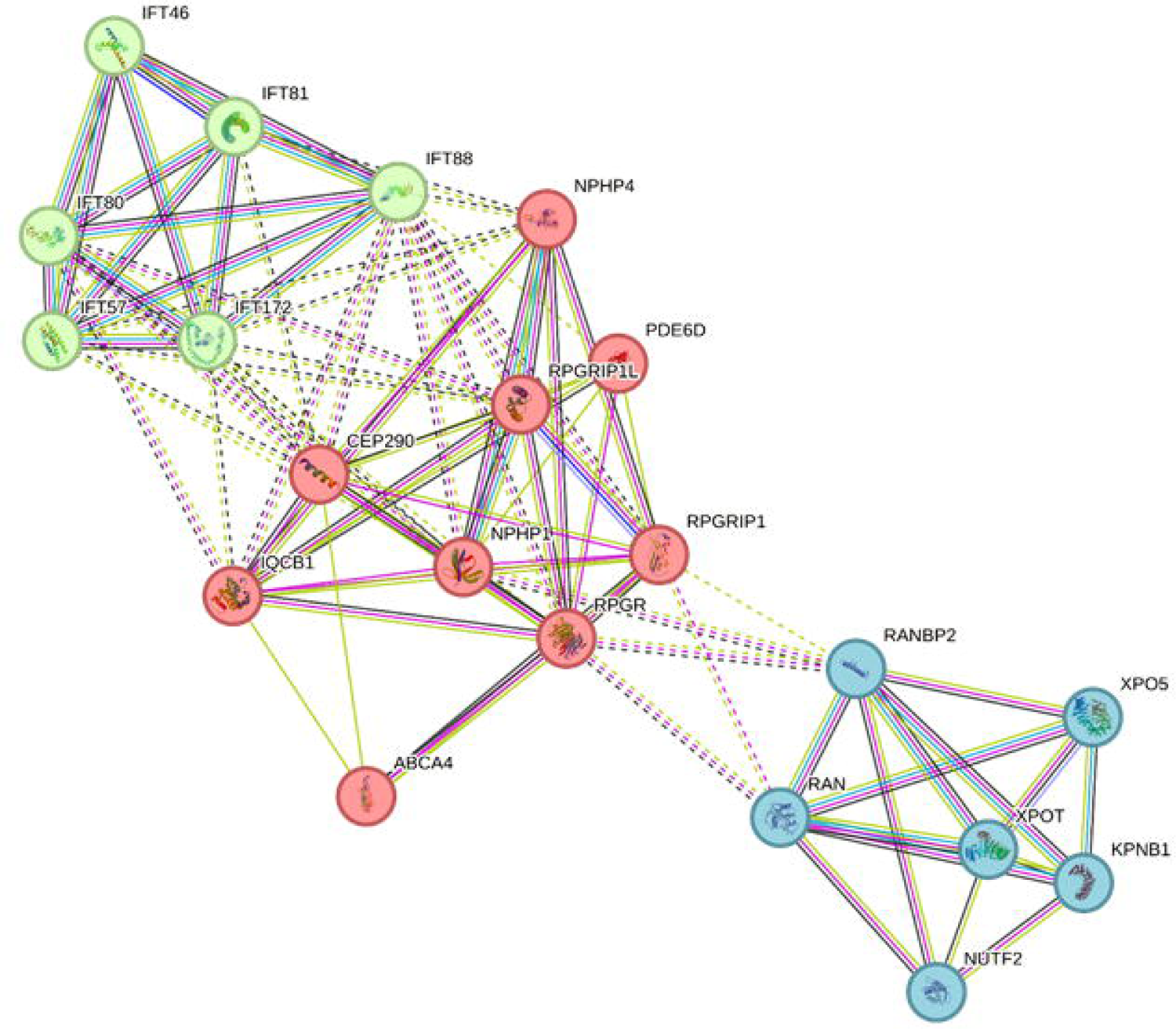
Protein–protein interaction analysis revealed the top RPGR-related proteins in three clusters. The red indicates the first cluster, the green indicates the second cluster, and the blue shows the third cluster of this interaction network. The first cluster modulates the cilium assembly pathway, the second regulates the chaperone-mediated autophagy pathway, and the third regulates the microRNA biogenesis.

**Table 1:**
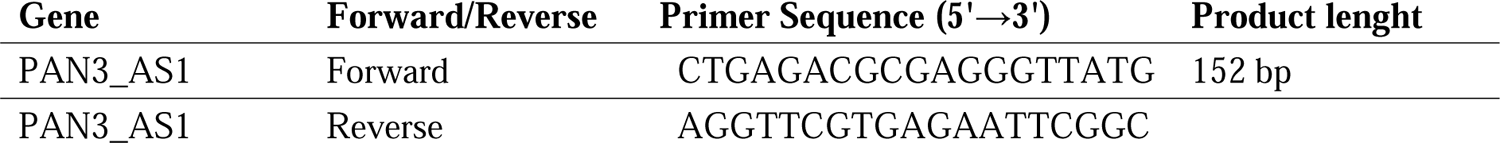
PAN3_AS1 Primer sequnces used for qRT-PCR experiment.

**Table 2:** lncRNA-mRNA interaction analysis. Top 10 targets of PAN3-AS1 are showed in this table.

| Sum of Energy | Min of Energy | Transcript ID | Transcript Name |
| --- | --- | --- | --- |
| -2,576.23 | -53.79 | ENST00000378505 | RPGR |
| -1,879.94 | -32.33 | ENST00000621744 | NBPF19 |
| -1,795.50 | -38.03 | ENST00000334976 | EMC10 |
| -1,745.36 | -35.45 | ENST00000622895 | GOLGA6L22 |
| -1,742.88 | -32.93 | ENST00000583866 | NBPF10 |
| -1,576.39 | -31.47 | ENST00000567107 | GOLGA6L2 |
| -1,429.80 | -34.15 | ENST00000619213 | GOLGA6L6 |
| -1,356.28 | -32.87 | ENST00000619423 | NBPF14 |
| -1,126.85 | -34.18 | ENST00000614055 | GOLGA6L1 |
| -272.13 | -26.7 | ENST00000400385 | AATBC |

**Table 3:** top 10 miRNAs regulating RPGR. miR-939-5p has the strongest interaction with RPGR mRNA.

| miRNA | Gene symbol | score | energy | seed | position |
| --- | --- | --- | --- | --- | --- |
| **hsa-miR-939-5p** | **RPGR** | **1** | **-29.3** | **1** | **3UTR** |
| hsa-miR-7160-5p | RPGR | 1 | -27.7 | 1 | 3UTR |
| hsa-miR-103a-1-5p | RPGR | 1 | -25.5 | 1 | 3UTR |
| hsa-miR-4496 | RPGR | 1 | -24.8 | 1 | 3UTR |
| hsa-miR-6846-5p | RPGR | 1 | -24.7 | 1 | 3UTR |
| hsa-miR-4277 | RPGR | 1 | -24.5 | 1 | 3UTR |
| hsa-miR-6757-5p | RPGR | 1 | -24.2 | 1 | 3UTR |
| hsa-miR-33b-3p | RPGR | 1 | -24 | 1 | 3UTR |
| hsa-miR-3670 | RPGR | 1 | -23.9 | 1 | 3UTR |
| hsa-miR-11401 | RPGR | 1 | -23.9 | 1 | 3UTR |

**Table 4:**
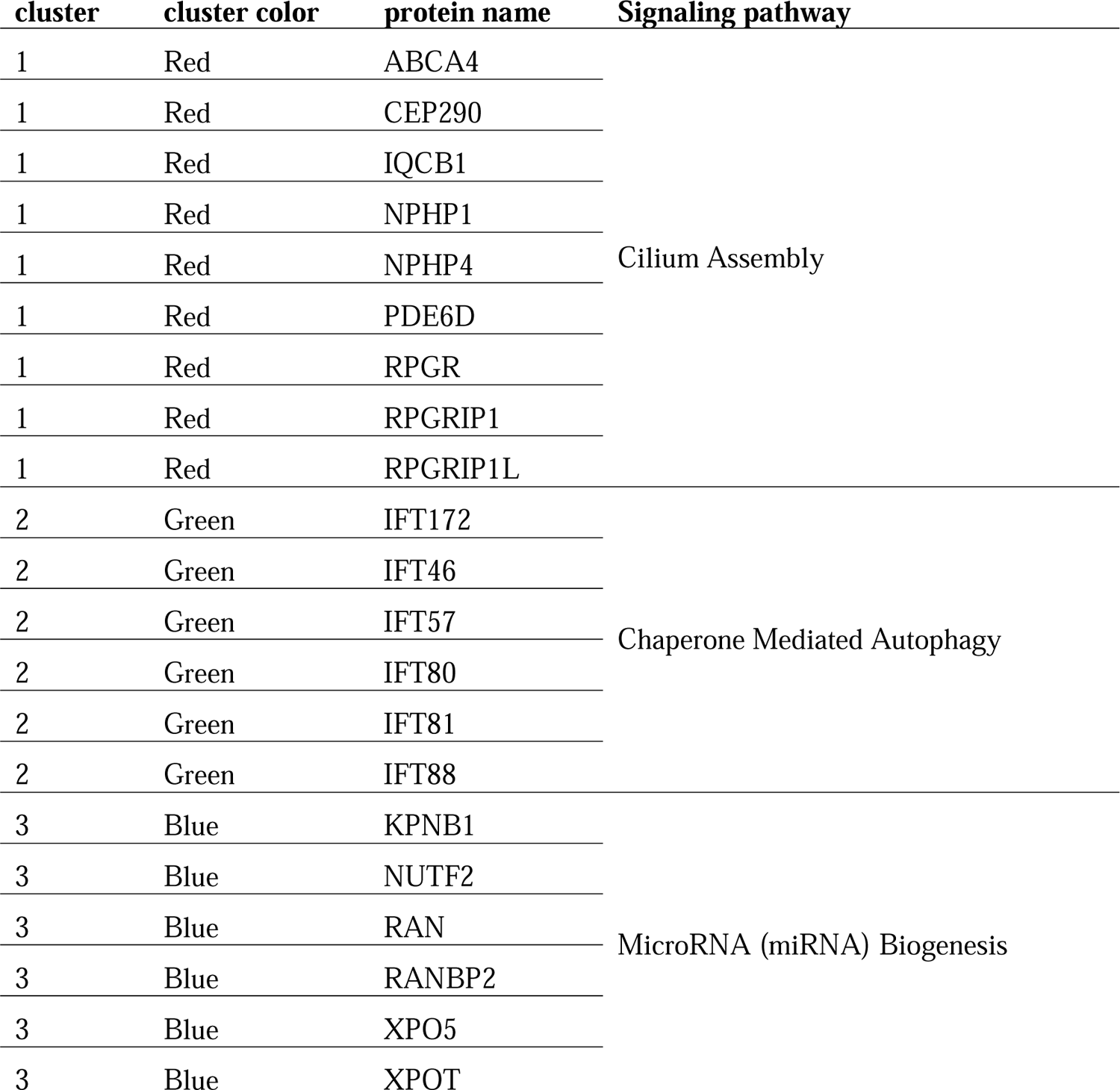
Protein interaction analysis table. Details of the cluster and those related signaling pathways are provided in the table.

| cluster | cluster color | protein name | Signaling pathway |
| --- | --- | --- | --- |
| 1 | Red | ABCA4 | Cilium Assembly |
| 1 | Red | CEP290 |  |
| 1 | Red | IQCB1 |  |
| 1 | Red | NPHP1 |  |
| 1 | Red | NPHP4 |  |
| 1 | Red | PDE6D |  |
| 1 | Red | RPGR |  |
| 1 | Red | RPGRIP1 |  |
| 1 | Red | RPGRIP1L |  |
| 2 | Green | IFT172 | Chaperone Mediated Autophagy |
| 2 | Green | IFT46 |  |
| 2 | Green | IFT57 |  |
| 2 | Green | IFT80 |  |
| 2 | Green | IFT81 |  |
| 2 | Green | IFT88 |  |
| 3 | Blue | KPNB1 | MicroRNA (miRNA) Biogenesis |
| 3 | Blue | NUTF2 |  |
| 3 | Blue | RAN |  |
| 3 | Blue | RANBP2 |  |
| 3 | Blue | XPO5 |  |
| 3 | Blue | XPOT |  |

Pathway enrichment analysis also revealed that the first cluster of the protein interaction network (which consisted of RPGR) modulates the cilium assembly signaling pathway (Figure 8). The second cluster regulates chaperone-mediated autophagy (Figure 9), and the third cluster has a significant role in the miRNA biogenesis. (Table 4).

**Figure 8:**
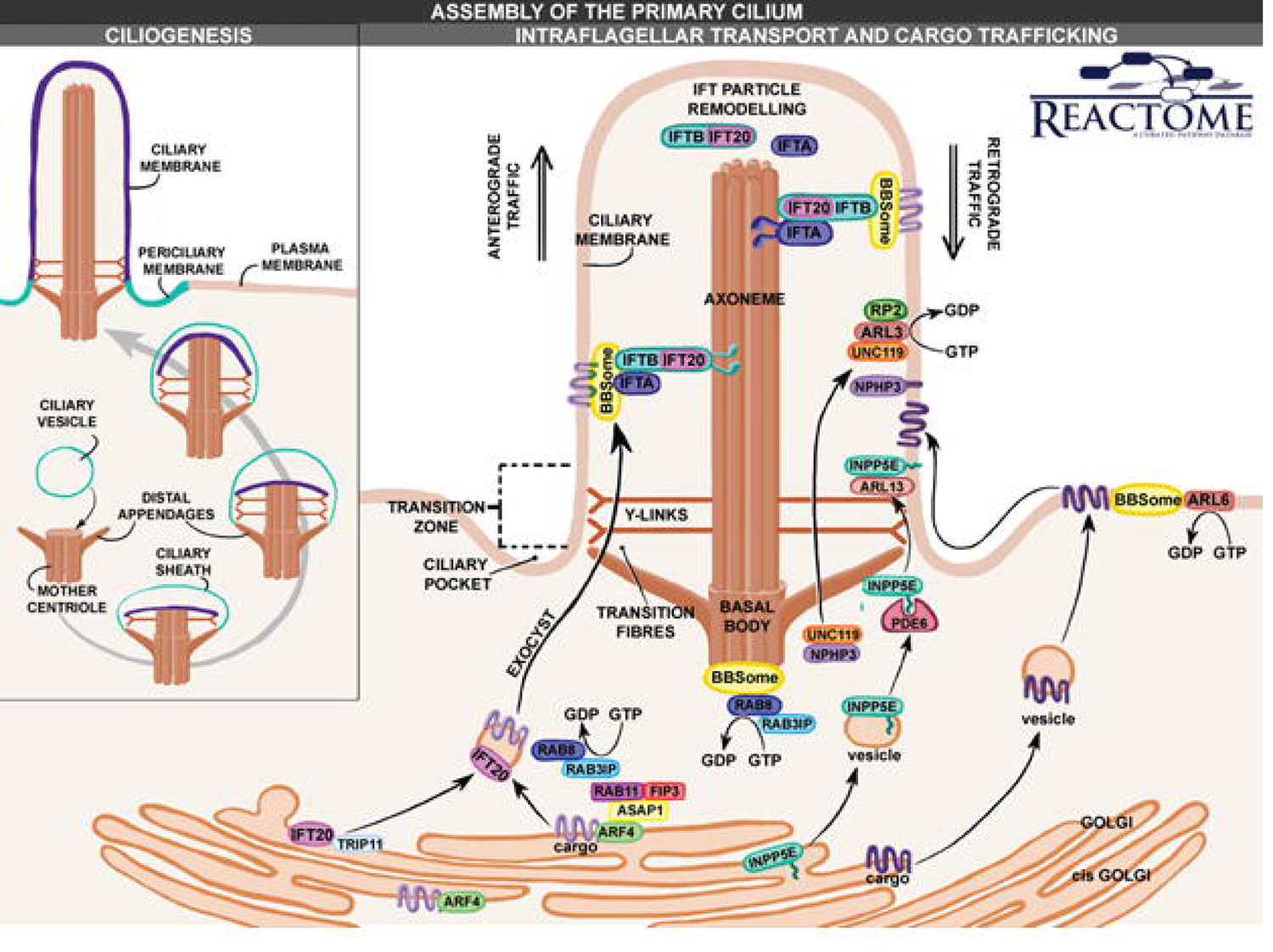
General overview of Cilium assembly signaling pathway.

**Figure 9:**
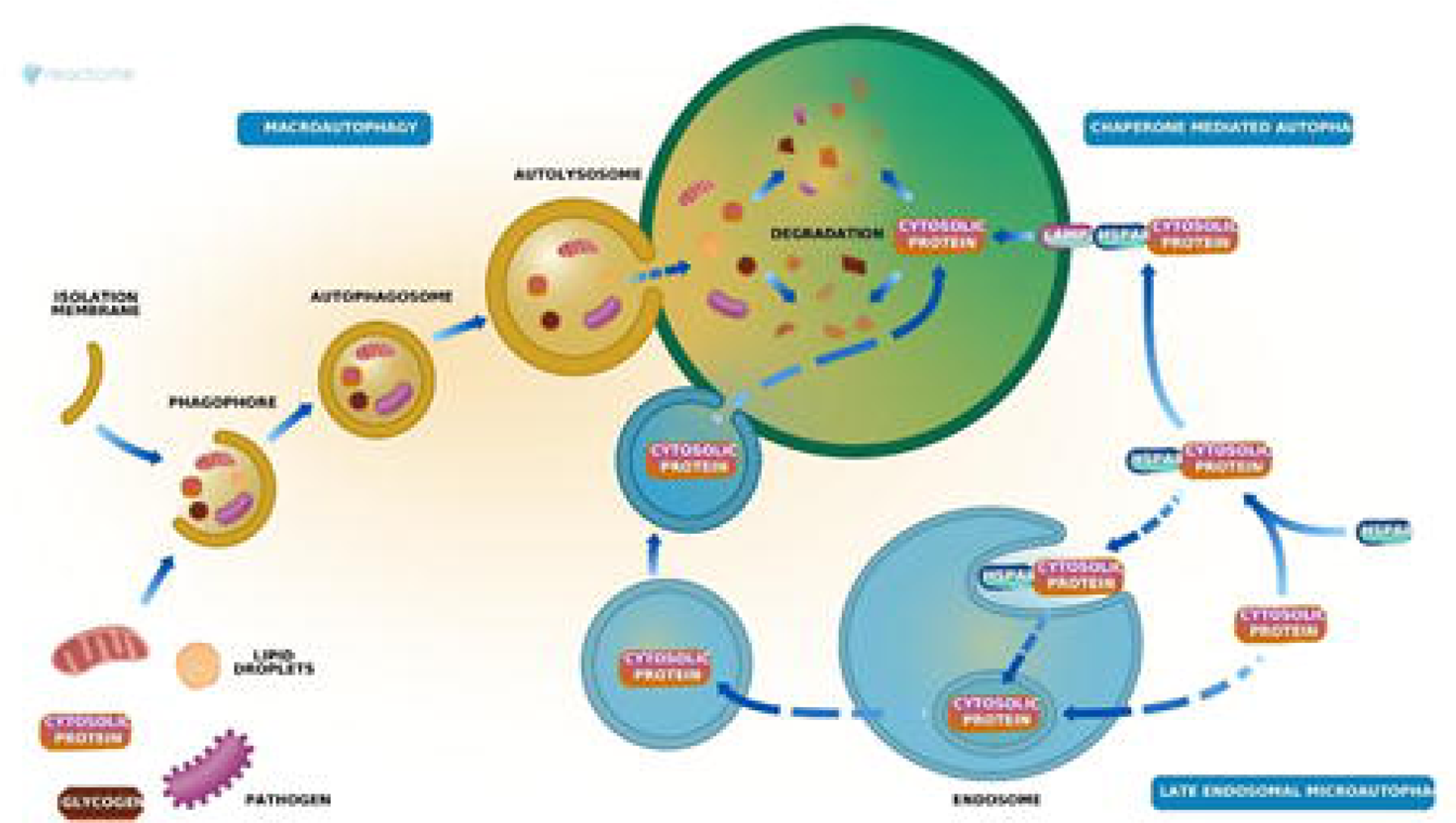
Schematic figure of autophagy signaling pathway.

## 4. Discussion

This study used an integrated systems biology approach to find a novel potential biomarkers of MS. Through the high-throughput gene expression, RNA and protein interaction, and pathway enrichment analysis, we found a novel lncRNA, PAN3-AS1, as a significant low-expressed potential biomarker in MS patients. Also, we discovered that PAN3-AS1 has a significant interaction with RPGR mRNA. Based on the protein interaction of RPGR, this protein has a substantial interaction with three protein groups. These proteins have been clusters based on those correlated signaling pathways. This interaction network regulates the following pathways: cilium assembly, chaperone-mediated autophagy, and miRNA biogenesis.

Furthermore, we find that miR-939-5p suppresses the expression level of RPGR through interaction with the 3’UTR region of its mRNA. There was no previous study about the gene expression pattern of PAN3-AS1 in MS patients. Also, this is the first study that mentioned the possible role of miR-939-5p in MS development.

Based on our investigations, cilium assembly is a crucial signaling pathway that might be regulated by PAN3-AS1 through the regulation of RPGR and its protein interactome. Based on the study of Mi Ki et al. in 2021 (Ki et al., 2021), Primary cilium in astrocytes is an essential neuronal signaling modulator in the central nervous system (CNS) (Ki et al., 2021). Also, based on this study, cilium in oligodendrocyte (OLG) regulates the myelination of CNS (Ki et al., 2021). Astrocytes, the most common glial cells in the CNS, are necessary for the creation and maintenance of the blood-brain barrier, as well as for the secretion of a range of neurotrophic factors during synaptogenesis, neuronal differentiation, and neuronal survival (Carson et al., 2006; Fakhoury, 2018). In neurodegenerative diseases, astrocytes produce harmful chemicals such as reactive oxygen species (ROS) and inflammatory mediators such as COX2, IL-1, TNF-, and IL-6 (Akiyama et al., 2000; McGeer & McGeer, 2002). When the CNS is destroyed, OPCs in the affected areas differentiate into OLGs. When this process malfunctions, less OLGs are created, resulting in MS development (Dulamea, 2017; Kuhn et al., 2019). As a result, earlier research has shown that differentiation of OLGs during neuronal protection/regeneration is critical in the treatment of MS (Baldassari et al., 2019; Stangel et al., 2017). As a result, various therapeutic potential targets have been presented, which are mostly involved in the formation or myelination of OLGs via Shh, Wnt, and LINGO1 signaling (Cole et al., 2017; Fancy et al., 2009; Ineichen et al., 2017).

Chaperone-mediated autophagy (CMA) is a kind of autophagy in which cellular components are delivered and degraded via a chaperone-dependent selective process. It is a catabolic process in lysosomes to maintain cellular homeostasis by lysis of excess or excessive soluble cytosolic proteins. It can be triggered in response to a number of stressors in order to restore cellular homeostasis (Li et al., 2018). CMA assists in the breakdown of various proteins that are prone to aggregate; hence, a decrease in CMA activity results in the formation of hazardous aggregates. CMA activity has been related to both an increase and a reduction in disease. In general, proteolytic mechanisms fail in neurodegenerative diseases, resulting in the buildup of harmful protein transformation (Andrade-Tomaz et al., 2020; Liao et al., 2021). Xu et al. 2021 define Metformin’s method of CMA induction as stimulation of TAK1-IKK signaling, which leads to phosphorylation of Ser85 of the central mediator of CMA, Hsc70, and its activation. The CMA substrate amyloid-beta precursor protein (APP) interacts with Hsc70 in an IKK/-dependent way. Inhibiting CMA-mediated APP degradation increases its cytotoxicity. Importantly, activation of CMA by Hsc70 overexpression or Metformin decreases accumulated brain A plaque levels and reverses the molecular and behavioral AD symptoms in the APP/PS1 animal model of Alzheimer’s disease (AD) (Xu et al., 2021). A growing number of studies have shown that autophagy is essential in the etiology of MS. Autophagy keeps the CNS balanced by digesting damaged organelles and aberrant proteins. Furthermore, autophagy plays a role in inflammatory responses by regulating immune cell activation and inflammatory factor release. However, the exact autophagy processes implicated in MS are not well-known (Shen et al., 2022).

The accurate regulatory mechanism of PAN3-AS1 and miR-939-5p still needs to be discovered. However, in this study, we demonstrated a possible correlation between the regulatory axis of PAN3-AS1/RPGR/miR-939-5p with the development of MS. It is highly suggested that the expression pattern of miR-939-5p and RPGR be evaluated in MS samples through more accurate experiments. Also, it is strongly recommended that the interaction of PAN3-AS1/RPGR/miR-939-5p can be evaluated through luciferase assay or RIP experiment.

## 5. Conclusion

lncRNA PAN3-AS1 could be a potential tumor suppressor and diagnostic biomarker of MS. This lncRNA could modulate cilium assembly and chaperone-mediated autophagy signaling pathway through regulation of RPGR mRNA transcription. RPGR and its protein interactome could be regulated by miR-939-5p.

## Data Availability

All data produced in the present work are contained in the manuscript

## Acknowledgments

We express our gratitude to Zist Fanavari Novin Biotechnology Institute, Department of Biology, University of Isfahan, and Department of Biology University of Zabol for their support.

